# Emergence of SARS-CoV-2 Alpha (B.1.1.7) variant, infection rates, antibody seroconversion and seroprevalence rates in secondary school students and staff: active prospective surveillance, December 2020 to March 2021, England

**DOI:** 10.1101/2021.07.14.21260496

**Authors:** Shamez N. Ladhani, Georgina Ireland, Frances Baawuah, Joanne Beckmann, Ifeanyichukwu O Okike, Shazaad Ahmad, Joanna Garstang, Andrew J Brent, Bernadette Brent, Felicity Aiano, Zahin Amin-Chowdhury, Meaghan Kall, Ray Borrow, Ezra Linley, Maria Zambon, John Poh, Lenesha Warrener, Angie Lackenby, Joanna Ellis, Gayatri Amirthalingam, Kevin E Brown, Mary E Ramsay

## Abstract

**Background:** In England, the rapid spread of the SARS-Cov-2 Alpha (B.1.1.7) variant from November 2020 led to national lockdown, including school closures in January 2021. We assessed SARS-CoV-2 infection, seroprevalence and seroconversion in students and staff when secondary schools reopened in March 2021.

**Methods:** Public Health England initiated SARS-CoV-2 surveillance in 18 secondary schools across six regions in September 2020. Participants provided nasal swabs for RT-PCR and blood samples for SARS-CoV-2 antibodies at the beginning (September 2020) and end (December 2020) of the autumn term and at the start of the spring term (March 2021).

**Findings:** In March 2021, 1895 participants (1100 students, 795 staff) were tested; 5.6% (61/1094) students and 4.4% (35/792) staff had laboratory-confirmed SARS-CoV-2 infection between December 2020 and March 2021. Nucleoprotein antibody seroprevalence was 36.3% (370/1018) in students and 31.9% (245/769) in staff, while spike protein antibody prevalence was 39.5% (402/1018) and 59.8% (459/769), respectively, similar to regional community seroprevalence. Between December 2020 and March 2021 (median 15.9 weeks), 14.8% (97/656; 95% CI: 12.2-17.7) students and 10.0% (59/590; 95% CI: 7.7-12.7) staff seroconverted. Weekly seroconversion rates were similar from September to December 2020 (8.0/1000) and from December 2020 to March 2021 (7.9/1000; students: 9.3/1,000; staff: 6.3/1,000).

**Interpretation:** By March 2021, a third of secondary school students and staff had serological evidence of prior infection based on N-antibody seropositivity, and an additional third of staff had evidence of vaccine-induced immunity based on S-antibody seropositivity. Further studies are needed to assess the impact of the Delta variant.

**Research in Context:** *Evidence Before this study:* The Alpha variant is 30-70% more transmissible than previously circulating SARS-CoV-2 strains in adults and children. One outbreak investigation in childcare settings estimated similar secondary attack rates with the Alpha variant in children and adults. There are limited data on the impact of the Alpha variant in educational settings. In England, cases in primary and secondary school aged children increased rapidly from late November 2020 and peaked at the end of December 2020, leading to national lockdown including school closures.

*Added Value of This Study:* Seroconversion rates in staff and students during December 2020 to March 2021, when the Alpha variant was the primary circulating strain in England, were similar to the period between September 2020 and December 2020 when schools were fully open for in-person teaching. By March 2021, a third of students overall and more than half the students in some regions were seropositive for SARS-CoV-2 antibodies. Among staff, too, around a third had evidence of prior infection on serological testing and a further third had vaccine-induced immunity.

*Implications of all the Available Evidence:* SARS-CoV-2 antibody seroprevalence was high among secondary school students in March 2021 and is likely to be higher following the emergence of an even more transmissible Delta variant in May 2021. Education staff are increasingly being protected by the national COVID-19 immunisation programme. These findings have important implications for countries that are considering vaccination of children to control the pandemic

## Introduction

In England, the emergence and rapid spread of the SARS-CoV-2 Alpha (B.1.1.7 or Kent) variant in November 2020 led to a second wave of cases, hospitalisations and deaths, resulting in prolonged national lockdown including school closures.(1-3) Following exposure to the virus, children are more likely to be asymptomatic or develop mild, transient and self-limiting upper respiratory tract infection compared to adults, who are more likely to develop severe disease, require hospitalisation and die of COVID-19. (4, 5)(6)

Early in the pandemic, schools in England were closed as part of national lockdown in March 2020 and only partially reopened for some school years in June 2020.(7) Since September 2020, however, all schools fully reopened for in-person teaching.(8) As per national guidance, face masks and face coverings were not recommended in classrooms, but staff and children in secondary schools were advised to wear them in communal areas outside the classroom if physical distancing was difficult to maintain.(8) Cases in adults and children increased throughout September and October 2020 and a second national lockdown was imposed for adults from 05 November to 02 December 2020, whilst keeping all schools open.(9) Cases fell rapidly first in adults and then in children even though all schools remained fully open at the time.(10)

To better understand infection and transmission of SARS-CoV-2 in secondary schools, Public Health England (PHE) initiated active, prospective surveillance in 18 schools across six English regions to assess the risk of SARS-CoV-2 infection in students and staff in September 2020.(11) In addition to nasal swabbing to identify acute SARS-CoV-2 infection, we also took blood samples from staff and students to measure SARS-CoV-2 antibody positivity, because it captures both symptomatic and asymptomatic prior infections, which are especially common in children.(11-13)

In December 2020, we found similar antibody seroprevalence and seroconversion rates in staff and students, which were comparable to local community rates at the time.(11) However, towards the end of November 2020, rapid increases in SARS-CoV-2 infection rates were observed across all age groups because of rapid increases in the more transmissible Alpha variant, leading to local tier restriction in areas of high community infections from 19 December 2020 and another national lockdown from 06 January 2021, including school closures from the end of the end of the autumn term in late December 2020 until 08 March 2021. Between 05 January and 08 March 2021 schools were closed for all students except vulnerable children or children of key workers and an estimated 5% of secondary school students and 25% of primary school students attended school during the lockdown period.(14) To assess the impact of the Alpha variant in students and staff, we undertook another round of nasal swabbing and blood sampling when participating schools reopened after prolonged national lockdown in March 2021, with the aim of assessing SARS-CoV-2 infection, antibody seroprevalence and seroconversion rates during a period of rapid spread of the Alpha variant in England.

## Methods

The COVID-19 Surveillance in Secondary School KIDs (sKIDsPLUS) protocol is available online (https://www.gov.uk/guidance/covid-19-paediatric-surveillance),(15) and results for the first two rounds of testing have been published.(11) The protocol was approved by PHE Research Ethics Governance Group (reference Nr0228; 24 August 2020). The study involved testing secondary school students for SARS-CoV-2 infection and antibodies at the start (Round 1: 22 September-17 October) and end (Round 2: 3-17 December) of the autumn term of the 2020/21 academic year and when the schools reopened in March (Round 3: 23 March-21 April)(**Supplementary Figure 1**). Secondary schools were approached in areas where a paediatric investigation team could be assembled: Derbyshire, West London, East London, Greater Manchester, Hertfordshire and Birmingham. Headteachers in participating schools emailed the study information pack to staff, parents of students aged <16 years and to students aged ≥16. Participants or their parent/guardian provided informed consent online via SnapSurvey, and completed a short questionnaire prior to the sampling day. The questionnaire requested information about demographics, risk factors and COVID-19 symptoms or confirmed infections in the household. Enrolment was open for new participants between Rounds 1 and 2, although 94 participants didn’t participate until round 3. A team of clinicians, nurses, phlebotomists and administrative staff attended the school on the sampling days. Local anaesthetic cream was offered to all students before blood sampling. A member of the school staff was present with each student. Participating students and staff had a nasal swab and a blood sample taken by the investigation team.

**Figure 1:**
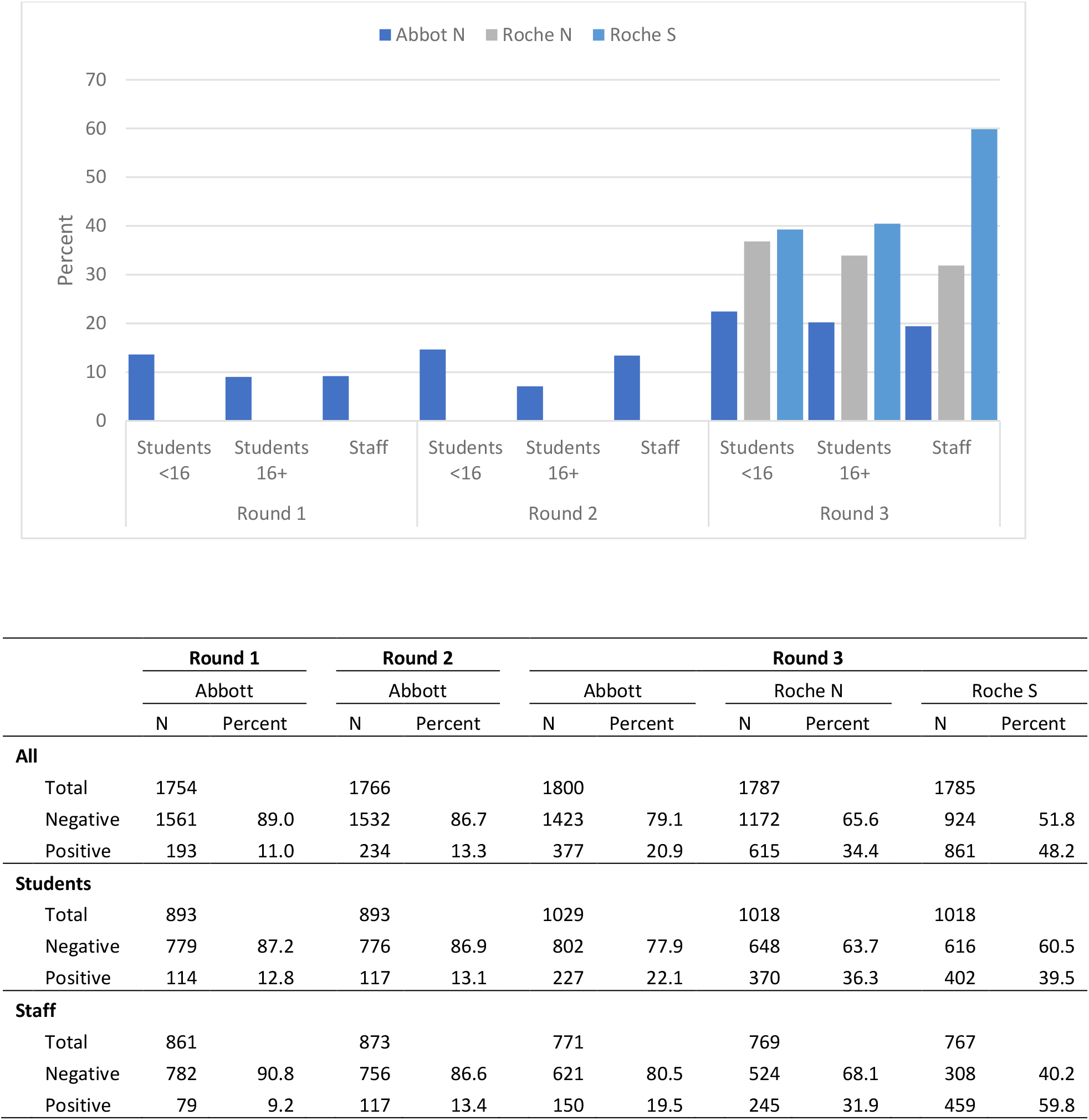
Seroprevalence in student and staff sKIDsPLUS participants using the Abbot N, Roche N and Roche S assays in Round 1 (September 2020), Round 2 (November/December 2020) and Round 3 (March/April 2021).

### Laboratory testing

The swabs were tested by a triplex reverse transcription PCR (RT-PCR) assay for the detection of ORF1ab and E gene regions of SARS-CoV-2 with simultaneous detection of an exogenous internal control using the Applied Biosystems Quantstudio 7-flex thermocycler (ThermoFisher Scientific, UK). The ORF1ab gene primers/probes published by the China CDC were combined with the E gene primers/probe published Corman *et al*., 2020.(16, 17) A positive RT-PCR result was reported to the participant, local investigator, head teacher and local PHE health protection team (HPT), typically within 48 hours of the sample being taken. The participant and household members self-isolated as per national guidance. Public health risk assessment was undertaken with the school to decide additional measures, including identification and isolation of the participant’s contacts inside and outside school premises.

Serology was performed on the Abbott Architect using a chemiluminescent microparticle immunoglobulin G (IgG) immunoassay targeting the nucleoprotein (N) (SARS-CoV-2 IgG, Abbott Commerce Chicago, USA) with a seropositivity threshold of 0.8 (henceforth referred to as Abbott N assay).(18) This assay was shown to detect SARS-CoV-2 N-antibodies as early as 7 days post symptom onset and is, therefore, particularly useful for assessing SARS-CoV-2 antibody seroconversion (negative to positive antibodies) between testing rounds.(19) Where sufficient serum was available, samples from Rounds 2 and 3 were additionally tested for nucleoprotein and spike (S) protein antibodies on the Roche Elecsys Anti-SARS-CoV-2 assay and Elecsys Anti-SARS-CoV-2 S assay respectively (henceforth referred to as Roche N and Roche S assays respectively).

### Statistical Analysis

Data were managed in R-Studio and Microsoft Access and analysed in Stata SE (version 15.1). Data that did not follow a normal distribution are described as median with interquartile ranges. Categorical data are described as proportions and compared with the Chi^2^-test or Fisher’s exact.

Participants were linked to routine laboratory report of SARS-CoV-2 to identify diagnoses between testing rounds. Firstly, NHS number was ascertained through linkage of participants to the Personal Demographics Service. A combination of NHS number, full name, sex, date of birth and postcode of residence was used to link participants to the routine laboratory reports of SARS-CoV-2 at PHE.

SARS-CoV-2 infection rate and antibody seroprevalence, with 95% confidence intervals (CI), were compared between secondary school students and staff. For comparisons with regional seroprevalence, three-week (round 3: 15 March-11 April) average prevalence data from PHE and NHS Blood and Transplant (NHS BT) serosurveillance of blood donors was provided. A sample of donors are tested for both N and S-antibodies on the Roche assays and for students data was compared to the seroprevalence in 18-30 year olds and staff were compared to 18-64 year old donors. Non-overlapping 95% CIs were used to assess statistical significance between student or staff rates and regional estimates. Antibody seroconversion rates with 95% confidence intervals were calculated for participants who were tested in two sequential rounds and were negative in their first round of testing.

Reported SARs-CoV-2 cases were used in conjunction with questionnaire data to ascertain likely time of infection. Reported SARS-CoV-2 diagnosis date was preferentially taken over reported date of symptom onset and reported date of diagnosis, as provided in the questionnaire.

Participants were classified as included in each round if they provided a blood or swab sample in that round. For SARS-CoV-2 antibody seroconversion, a multivariable regression model was built using age, sex, ethnicity, school area and school attendance during January to March 2021 lockdown. Variation between schools was allowed for via a school-level random effect.

## Results

Eighteen secondary schools in six school areas enrolled and participated in three rounds of sKIDsPLUS testing. Overall, 1895 participants were tested in Round 3, 1100 students and 795 staff (**Table 1**). Of those who completed the online surveillance questionnaire for Round 3 (students: 78.4%; n=862, staff: 88.4%; n=703), most (82.8%; 707/854) students had not attended school during the third lockdown, with 5.7% (n=49) attending part-time and 11.5% (n=98) attending full-time. In comparison, the staff had all attended school, either part-time (70.3%; 352/501) or full-time (29.7%; n=149). Two-fifths (42.9%; 300/700) of staff were vaccinated, and 1.3% (11/852) students reported being vaccinated).

**Table 1:**
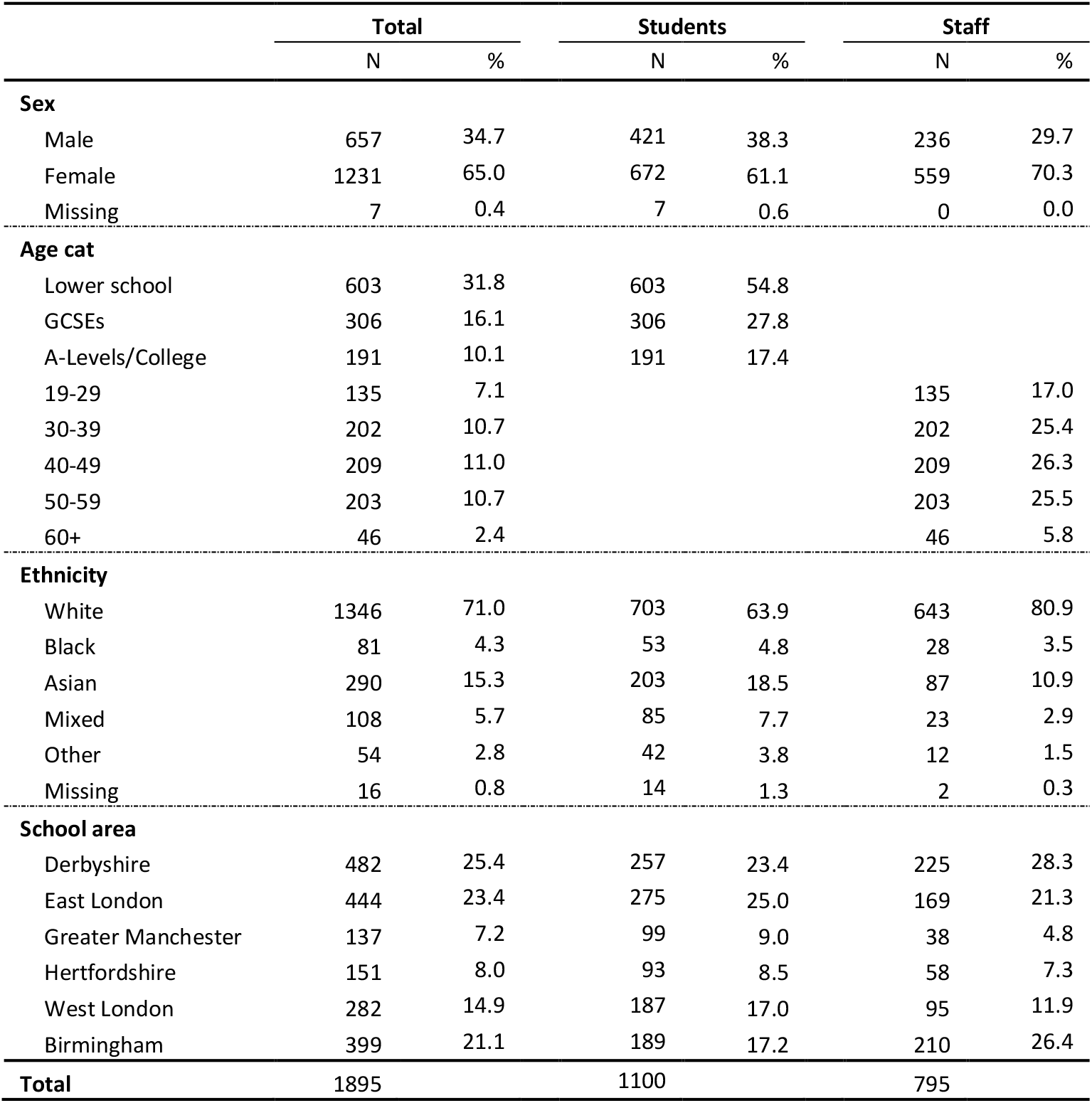
Characteristics of students and staff from 18 secondary schools participating in sKIDs PLUS in Round 3 (March-April).

### RT-PCR Results

In March 2021 (Round 3), two (0.18%; n=1,094) students from different schools, and none of the 792 staff tested were SARS-CoV-2 RT-PCR positive on nasal swabs. For the ORF1AB gene, one swab had a cycle threshold (Ct) value of 21.9 and the other was 38.1, for the E-gene the corresponding Ct-values were 20.5 and 32.6. Sequencing was successful for the swab with the lower ORF1AB and E gene Ct values and was identified as the Alpha (B.1.1.7) variant. Linkage to the national SARS-CoV-2 testing database identified an additional 95 cases, including 60 in students and 35 in staff. This equates to a diagnosed infection rate of 5.7% (62/1094) in students and 4.4% (35/792) in staff between Rounds 2 and 3.

### Antibody prevalence

Serum samples were tested on the Abbott N-antibody platform for all three rounds and seroprevalence increased from 11.0% (193/1754) in Round 1, 13.3% (234/1766) in Round 2 to 20.9% (377/1800) in Round 3 (p<0.001 for both rounds vs. Round 3) (**Figure 1**). Because of an assay-related high antibody waning rate over time with the Abbott N-antibody assay, sera from Round 3 were also tested on the Roche N and S assay. Using the Roche assay, the N-antibody seroprevalence was 36.3% (370/1018) in students and 31.9% (245/769) in staff, while S-antibody prevalence was 39.5% (402/1018) and 59.8% (459/767), respectively (**Figure 2**). In S-antibody positive staff, 63.0% (259/411) reporting being vaccinated.

**Figure 2:**
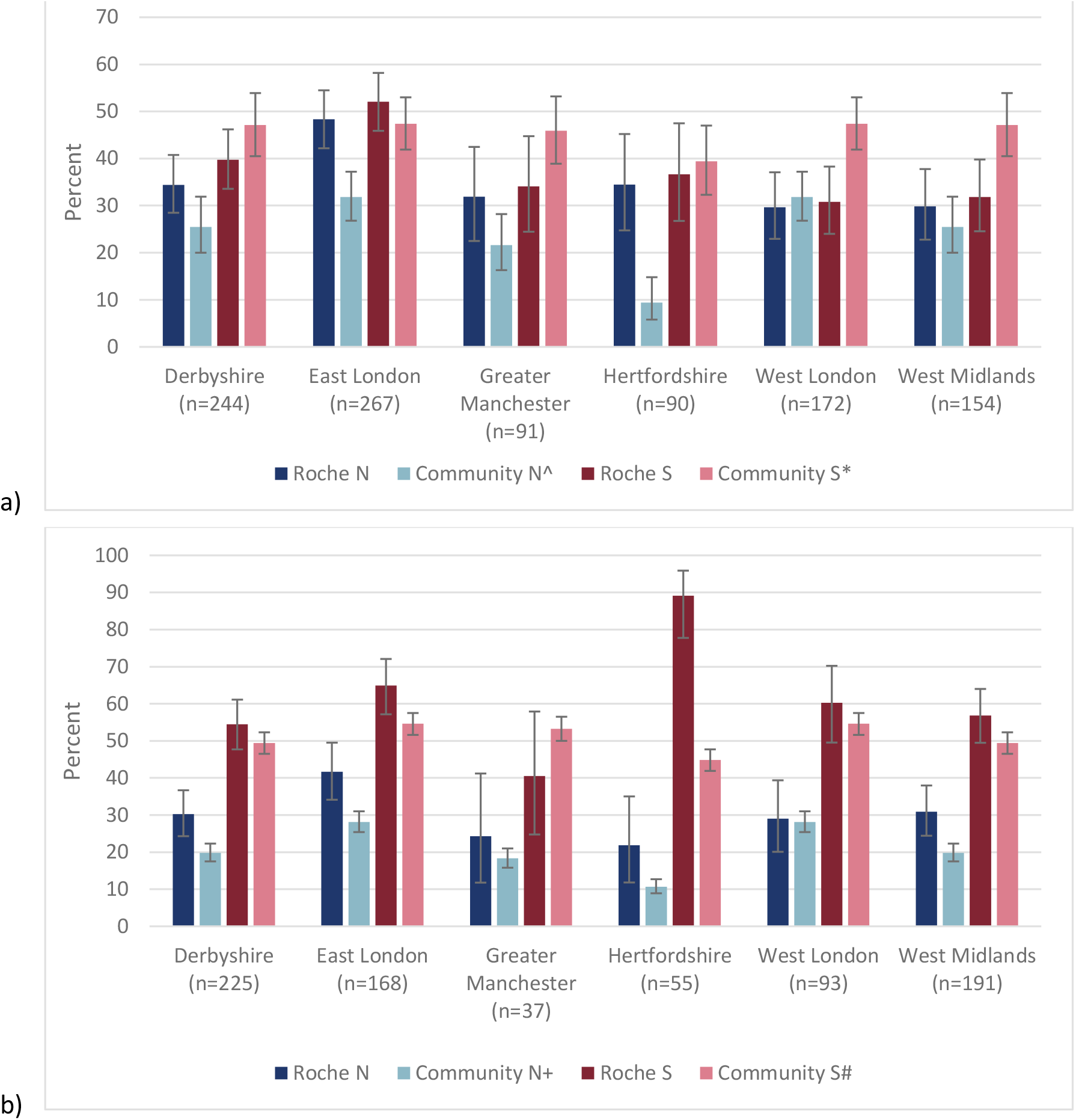
Seroprevalence in student (a) and staff (b) sKIDsPLUS participants by region using the Roche N and Roche S assays in Round 3 (March/April 2021) and compared to community estimates for N and S antibodies. *Source: NHSBT Seroprevalence data for the period 15*^*th*^ *March to 11*^*th*^ *April: * S antibody prevalence in 18-30 year olds by region; ^ N antibody prevalence in 18-30 year olds; # S antibody prevalence in 18-64 year olds by region; + N antibody prevalence in 18-64 year olds by region*.

When compared to regional seroprevalence data using the NHS BT samples, N-antibody seroprevalence was higher in students from East London and Hertfordshire, and higher in staff from Derbyshire, East London and West Midlands. For S-antibody, seroprevalence was higher in students from West London and West Midlands, and higher in staff from Hertfordshire (**Figure 2**).

### Seroconversion rates

Overall, 758 students and 688 staff had Abbott N-antibody results from both Rounds 2 and 3, including 656 (86.5%) and 590 (85.8%), respectively, who were negative in Round 2. Between Rounds 2 and 3, 14.8% (97/656; 95% CI: 12.2-17.7) of students and 10.0% (59/590; 95% CI: 7.7-12.7) of staff seroconverted over a median of 15.9 weeks (IQR: 15.1-15.9) (**Table 2**). Seroconversion rate was 9.3 per 1,000 weeks (95% CI: 7.5-11.3) between Rounds 2 and 3 in students, compared to 6.8 per 1,000 weeks (95% CI: 4.8-9.4) between Rounds 1 and 2. Among staff, the seroconversion rate was 6.3 per 1,000 weeks (95% CI: 4.8-8.1) between Rounds 2 and 3 and 8.9 per 1,000 weeks (95% CI: 6.7-11.5) between Rounds 1 and 2

**Table 2:** Seroconversion percent and rate per 1,000 weeks for sKIDs PLUS students and staff between Rounds 1 and 2 and 2 and 3, including by frequency of attendance. *\*breakdown of attendance will not add to student and staff totals as questionnaire not returned by all participants*

|  | Sample | Seroconverters |  |  |  |
| --- | --- | --- | --- | --- | --- |
|  |  | N | Percent | Rate per 1,000 weeks | 95% CI |
| Round 1->2 |  |  |  |  |  |
| Total | 1201 | 93 | 7.7 | 8.0 | 6.4-9.7 |
| Students | 549 | 36 | 6.6 | 6.8 | 4.8-9.4 |
| Staff | 652 | 57 | 8.7 | 8.9 | 6.7-11.5 |
| Round 2->3 |  |  |  |  |  |
| Total | 1246 | 156 | 12.5 | 7.9 | 6.7-9.2 |
| Students | 656 | 97 | 14.8 | 9.3 | 7.5-11.3 |
| *Not attending school* | 432 | 44 | 10.2 | 6.3 | 4.6-8.5 |
| *Attending school part time* | 33 | 8 | 24.2 | 15.6 | 6.8-30.5 |
| *Attending school full time* | 54 | 13 | 24.1 | 15.5 | 8.3-26.4 |
| Staff | 590 | 59 | 10.0 | 6.3 | 4.8-8.1 |
| *Not attending school* | 0 | - |  |  |  |
| *Attending school part time* | 265 | 25 | 9.4 | 6.0 | 3.9-8.8 |
| *Attending school full time* | 103 | 16 | 15.5 | 9.9 | 5.7-16.0 |

Where known, more student seroconverters were asymptomatic than staff (66.7%; 44/66 vs. 30.9%; 17/55; p<0.001). When using linked national testing data and reported symptoms and positive test dates in the period between Rounds 2 and 3 to estimate likely date of infection in those who seroconverted, half (56.5%; 48/85) of the cases were in the last 4 weeks of 2020, 28.2% (24/85) were in the first 4 weeks of 2021 and 12.9% (11/85) were in the weeks 5-8 of 2021 (**Supplementary table 2**).

Using a logistic regression, seroconversion in students between Rounds 2 and 3 was associated with school area (p=0.0060), with higher rates in East London (adjusted Odds Ratio [OR]: 3.60; 95% CI: 1.59-8.14 compared to Derbyshire) and full-time school attendance during the third lockdown (OR: 2.27; 95% CI: 1.06-4.86) when compared to students studying at home (**Table 3**). In staff, seroconversion was associated with Asian ethnicity (OR: 2.95; 95% CI: 1.11-7.84) when compared to White participants. Clustering was not significant at a school level for students (p=1.0) or staff (p=0.068).

**Table 3:** Risk factors for seroconversion in students (n=511) and staff (n=358) between Round 2 (December 2020) and Round 3(March 2021).

|  | Students (clustering p=1.0) |  | Staff (clustering p=0.068) |  |
| --- | --- | --- | --- | --- |
|  | adjusted OR (95% CI) | p-value | adjusted OR (95% CI) | p-value |
| **Sex** |  |  |  |  |
| Male | 1.46 (0.78-2.74) | 0.24 | 1.37 (0.66-2.86) | 0.40 |
| Female | 1 |  | 1 |  |
| **Age cat** |  |  |  |  |
| Lower school | 1 |  |  |  |
| GCSEs | 1.04 (0.51-2.13) | 0.97 |  |  |
| A-Levels/College | 1.11 (0.52-2.38) |  |  |  |
| 19-29 |  |  | 0.42 (0.11-1.67) |  |
| 30-39 |  |  | 1 |  |
| 40-49 |  |  | 0.69 (0.26-1.81) | 0.79 |
| 50-59 |  |  | 0.76 (0.31-1.88) |  |
| 60+ |  |  | 0.70 (0.17-2.90) |  |
| **Ethnicity** |  |  |  |  |
| White | 1 |  | 1 |  |
| Black | 2.33 (0.63-8.69) | 0.71 | - | 0.083 |
| Asian | 1.30 (0.61-2.75) |  | 2.95 (1.11-7.84) |  |
| Mixed | 1.06 (0.33-3.38) |  | 0.70 (0.08-6.16) |  |
| Other | 0.72 (0.15-3.50) |  | - |  |
| **School area** |  |  |  |  |
| Derbyshire | 1 |  | 1 |  |
| East London | 3.60 (1.59-8.14) |  | 1.10 (0.32-3.76) |  |
| Greater Manchester | 1.21 (0.29-5.06) | 0.0060 | 0.29 (0.03-3.15) | 0.69 |
| Hertfordshire | 0.96 (0.23-3.99) |  | 0.92 (0.13-6.69) |  |
| West London | 1.49 (0.54-4.12) |  | 0.35 (0.07-1.72) |  |
| Birmingham | 0.66 (0.18-2.36) |  | 0.73 (0.23-2.33) |  |
| **School attendance during lockdown** |  |  |  |  |
| Not attending school | 1 |  |  |  |
| Attending school part time | 2.52 (0.93-6.81) | 0.032 | 1 | 0.066 |
| Attending school full time | 2.27 (1.06-4.86) |  | 2.02 (0.95-4.28) |  |

## Discussion

Between December 2020 and March 2021, 5.7% of secondary school students and 4.4% of staff had laboratory-confirmed COVID-19, mainly during December 2020, when schools were open and case rates of the SARS-CoV-2 Alpha variant were increasing rapidly across England. When assessed using serum antibodies, however, the estimated proportions exposed to the virus over the same period were 14.8% and 10.0%, respectively, likely because of asymptomatic and pauci-symptomatic infections that did not warrant testing for the virus. The weekly seroconversion rate in staff and students was similar between December 2020 and March 2021 when compared with September to December 2020, even though the majority students did not attend school and more than two-thirds of staff attended school part-time during the latter period. By March 2021, 36.3% in students and 31.9% in staff had evidence of SARS-CoV-2 infection based on N-antibody seropositivity. This was supported by similar S-antibody seropositivity of 39.5% in students who were unvaccinated at the time, but 59.8% of staff had detectable spike antibodies because two-thirds of the school staff had been vaccinated as part of the national COVID-19 vaccine rollout.

Serum antibody testing is the most robust investigation for assessing prior exposure to SARS-CoV-2 because it captures both symptomatic and asymptomatic infections, the latter contributing to a significant proportion of total cases among both students (67%) and staff (31%) in our cohort. This is consistent with the published literature, but due to testing and isolation guidance, PCR-testing only provides a point estimate of asymptomatic infection prevalence in participants who were in school on the day of testing.(20) At the same time, testing of symptomatic individuals only will miss more than half the infections, as evidenced through linkage of our cohort with the national SARS-CoV-2 testing database, which estimated an infection prevalence of around 5% compared to more than twice this estimate through seroconversion.

We initially used the Abbott N-antibody testing platform because our early validation studies indicated that seropositivity was detected quicker after acute infection compared to other commercial platforms.(19, 21, 22) Using this assay, we found that the weekly seroconversion rates were similar between September-December 2020 and from December 2020 to March 2021, despite most students not attending school and staff attending part-time only during the latter period. The date of PCR-testing among symptomatic participants indicated that most infections occurred during the peak of the Alpha variant outbreak nationally in December 2020, although cases also occurred at lower rates during January and February 2021. Notably, attending school was associated with an increased risk of seroconversion in students, and marginally non-significant in staff, although we were not able to ascertain whether the infection was acquired inside or outside school. Data from the Schools Infection Survey (SIS) also showed similar N-antibody seroconversion rates in among primary and secondary school staff for the autumn term and lockdown period, although data on students and more detailed estimates by attendance are yet to be published.(23) A number of epidemiological studies have reported that most confirmed SARS-CoV-2 infections in school-aged children are acquired outside school, usually from a household member, with very low rates of in-person transmission of SARS-CoV-2 within school premises, even with active case finding.(24-26) This is consistent with seroprevalence studies in both primary and secondary schools when all students were attending in-person teaching.(11, 12, 27) That seroconversion rates were lower in students who did not attend in-person teaching is not surprising since their risk of coming into contact with an infected person would be significantly reduced.

An as-yet unexplained phenomenon of the Abbott nucleoprotein assay is a very high antibody seroreversion rate over time.(21) This appears to be specific to this particular platform because such a high seroreversion rates is not observed when the same sera are tested on other commercial nucleoprotein or spike protein antibody platforms.(21) Consequently, we re-tested all samples in round 3 in the Roche N assay, which reported seroprevalence rates around 65% higher than the Abbott nucleoprotein assay (Figure 1).

By March 2021, a third of students had SARS-CoV-2 antibodies, ranging between 30% in West London to almost 50% in East London. In students, the antibodies were most likely through natural infection, since vaccination was not recommended for this age-group. Observational data from England indicates that secondary attack rates followed an inverted U relationship with age, such that the risk of transmission increased from 9% in 0-9 year-olds to 12% among 10-19 year-olds and 16% among 20-29 year-olds and remaining at 20-24% thereafter.(28) Additionally, secondary attack rates were higher across all age-groups for the Alpha variant compared to previously circulating strains.(3) An outbreak investigation in three German childcare centres found infection and transmission rates with the Alpha variant were similar for children and for adults but higher than previously circulating SARS-CoV-2 strains.(29) Increased transmission would explain the rapid increase in infection rates among secondary school students attending school during late November and December 2020, and the high seroconversion rates by March 2021, despite schools being closed for most of this period. Reassuringly, the severity of clinical disease in children with the B.1.1.7 variant was similar to previously circulating strains.(10, 30)

Since April 2021, the Alpha variant has been replaced by an even more transmissible Delta variant in England, and, although cases remained low when schools reopened full-time for in-person teaching on 08 March 2021, whilst adults remained in lockdown, the transition to the easing of national lockdown on 17 May was associated with an increase in cases among young adults and older teenagers nationally.(31, 32) Older adults, however, remain protected against infection, hospitalisation and deaths because of the national rollout of the COVID-19 immunisation programme since the end of December 2020.(32) The programme prioritised vaccination for older adults, followed by health and social care workers, those with underlying co-morbidities and the progressively in younger adults in 10-year age-bands.(33) School staff were not prioritised for vaccination in the UK because their risk of COVID-19 was similar to other professions.(34) Instead, they were vaccinated according to their age and comorbidity status. Since we also measured spike antibodies using the Roche S assay in Round 3, we were able to estimate that around 60% of school staff were protected by March 2021, either through natural infection or vaccination. Since 17 May, however, cases and outbreaks have been increasing in school-aged children, with the Delta variant now accounting for >90% of SARS-CoV-2 infections in England.(32, 35) It is likely that SARS-CoV-2 antibody prevalence in students will now be higher than in March 2021.

The high seroprevalence rate in secondary school-aged children has implications for vaccinating secondary school students following approval of the Pfizer-BioNTech mRNA vaccine from adults down to 12 year-olds.(36) Some countries including the United States, Canada and Israel have already started vaccinating adolescents but others, including the UK, are yet to make a recommendation. The decision to recommend COVID-19 vaccination for teenagers should consider the added value of vaccinating a group with a high prevalence of natural immunity or, alternatively, whether a single dose, or a lower dose, might be sufficient to confer immunity whilst limiting the risk of potential adverse events, including myocarditis in teenagers and young adults.(37-40) Reassuringly, from 18 June 2020, all school staff are now eligible for vaccination – the School Infection Survey recently reported that 85% of staff had received at least one vaccine dose by May 2021.(23, 41)

### Limitations

The strength of this surveillance lies in the rapid recruitment and longitudinal assessment of secondary schools as soon as they reopened after national lockdown in September 2020. There are, however, some limitations. We only recruited schools in regions where we had paediatric investigation teams to take blood samples from large numbers of staff and students. Our schools are, therefore, not intended to be representative of all secondary schools in England. We avoided recruiting some secondary school years with end-of-year national examinations to minimise disruption of their education. Additionally, as already discussed, we were unable to assess whether the confirmed infections occurred within or outside the school premises. We are currently conducting a larger national School Infection Survey (SIS) involving 150 schools across England during the 2020/21 academic year.(42)

## Conclusions

Following the emergence of the Alpha variant in the UK, a third of students and staff had evidence of prior SARS-CoV-2 infection by March 2021, despite national lockdown including school holidays and closures for almost three months. Antibody seroprevalence rates are likely to be substantially higher with the emergence and rapid spread of the more transmissible Delta variant associated with increasing rates of SARS-CoV-2 infections and COVID-19 outbreaks in educational settings since mid-May 2021. Most adults including education staff and household members, however, are now protected through the national COVID-19 immunisation programme. These findings are important when considering vaccination of teenagers against COVID-19.

## Data Availability

Applications for relevant anonymised data should be submitted to the Public Health England Office for Data Release.

https://www.gov.uk/government/publications/accessing-public-health-england-data/about-the-phe-odr-and-accessing-data

## Acknowledgements

The authors would like to thank the schools, headteachers, staff, families and their very brave children who took part in the sKIDsPLUS surveillance. The authors would also like to thank the staff in the Immunisation Department and in the Virus Reference Department (VRD) at PHE Colindale, as well as Sero-epidemiology Unit (SEU) at PHE Manchester for supporting the sKIDS surveillance.

## Contributors

SNL, FB, JB, IOO, SA, JG, AJB, BB, GA, VS, KEB, and MER were responsible for conceptualisation and study design and methodology. SNL, FB, JB, IOO, SA, JG, AJB, BB, GI, FA, ZA-C, RB, EL, MZ, AL, JE and JP contributed to project administration (including laboratory colleagues). SNL and GI contributed to the original draft of the report, did the formal analysis and were responsible for data validation and verification. All authors contributed to reviewing and editing of the manuscripts. All authors had access to the data; SNL and GI had final responsibility to submit for publication.

