## Supplementary Materials for "Emergence of SARS-CoV-2 Alpha (B.1.1.7) variant, infection rates, antibody seroconversion and seroprevalence rates in secondary school students and staff: active prospective surveillance, December 2020 to March 2021, England"

**Supplementary Results**

**Supplementary Figure 1: Flow of sKIDs PLUS student and staff participants between rounds 1, 2 and 3.**

*****signed up prior to rounds 1 and 2 but did not participate until round 3*

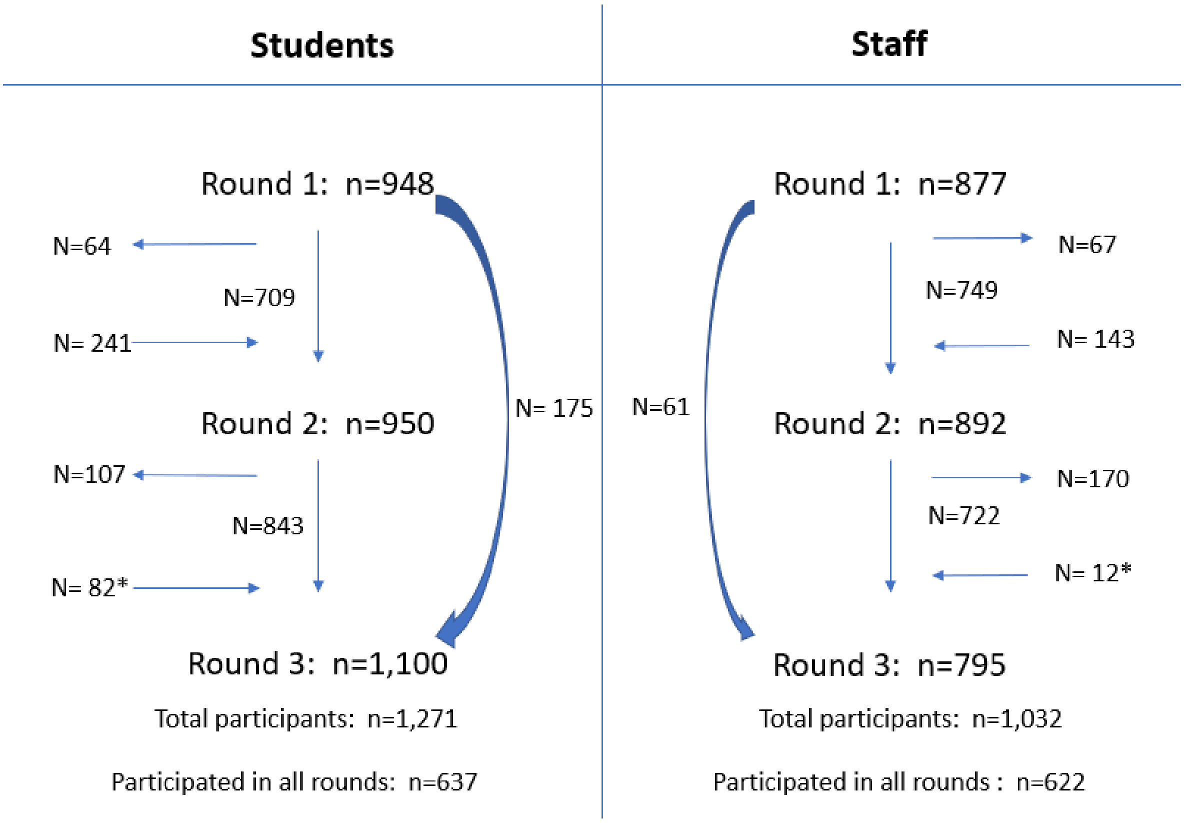

**Supplementary figure 2: The likely timing of SARS-CoV-2 infection* in 86 seroconverters, where information is available, between Rounds 2 and 3 compared to the number of positive cases in England for the corresponding weeks.**

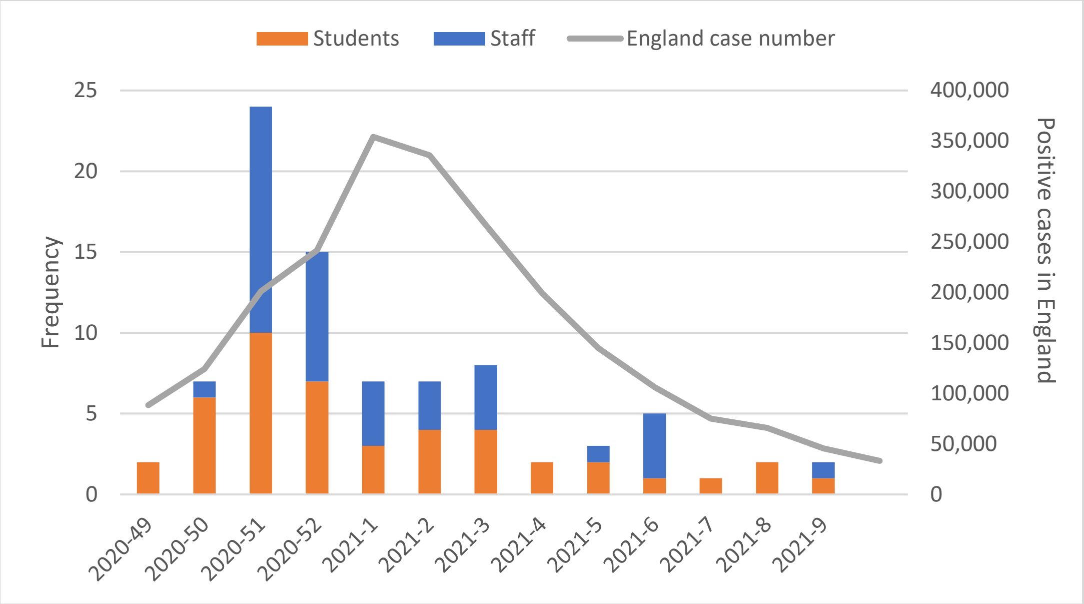


** Time of infection estimated using linked national testing records, reported positive tests and reported date of symptom onset in a hierarchy of national testing records, followed by a reported (via the round 3 questionnaire) positive test date and date of symptom onset*
